# Improved prediction of new COVID-19 cases using a simple vector autoregressive model: Evidence from seven New York State counties

**DOI:** 10.1101/2022.01.14.22269324

**Authors:** Takayoshi Kitaoka, Harutaka Takahashi

## Abstract

With the rapid spread of COVID-19, there is an urgent need for a framework to accurately predict COVID-19 transmission. Recent epidemiological studies have found that a prominent feature of COVID-19 is its ability to be transmitted before symptoms occur, which is generally not the case for seasonal influenza and SARS. Several COVID-19 predictive epidemiological models have been proposed; however, they share a common drawback—they are unable to capture the unique asymptomatic nature of COVID-19 transmission. Here, we propose vector autoregression (VAR) as an epidemiological county-level prediction model that captures this unique aspect of COVID-19 transmission by introducing newly infected cases in other counties as lagged explanatory variables. Using the number of new COVID-19 cases in seven New York State counties, we predicted new COVID-19 cases in the counties over the next 4 weeks. We then compared our prediction results with those of 11 other state-of-the-art prediction models proposed by leading research institutes and academic groups. The results showed that VAR prediction is superior to other epidemiological prediction models in terms of the root mean square error of prediction. Thus, we strongly recommend the simple VAR model as a framework to accurately predict COVID-19 transmission.

## 1. INTRODUCTION

Severe acute respiratory syndrome coronavirus 2 (SARS-CoV-2), identified in 2019, has caused the coronavirus disease 2019 (COVID-19) pandemic. With the rapid spread of COVID-19, there is an urgent need for a framework to accurately forecast COVID-19 progression. To this end, a variety of COVID-19 epidemiological forecasting models have been proposed by major research institutes. Wang et al. (2020) classified forecasting models into three categories: a) mechanistic models, b) time series models, and c) models based on deep learning. Examples of mechanistic models are the susceptible–infected– recovered (SIR) model and the modified susceptible–exposed–infected–recovered (SEIR) population propagation model. The majority of deep learning models extend mechanistic models with deep learning methods. In this study, we compared the forecasting accuracy of our model to that of 11 state-of-the-art forecasting models proposed by major research institutes and academic groups.

To predict the number of new COVID-19 cases by county, Shang et al. (2021) recently proposed a data-driven regression model called the vector autoregression (VAR) epidemiological model^1^. VAR is a time series model and contrasts with mechanistic and deep learning models in two aspects: 1) VAR solely uses county-level new COVID-19 cases as the forecasting data; and 2) VAR captures COVID-19 cross-county transmission by introducing other counties’ COVID-19 case data as lagged explanatory variables. The second point is important, because to predict COVID-19 cases at the county level, it is necessary to consider cross-county infection as a transmission mechanism of SARS-CoV-This is different from that of other viral infections such as seasonal influenza and SARS. To characterize the transmission dynamics of COVID-19, two important epidemiological terms were introduced: the incubation period (the time between infection and the onset of symptoms) and the serial interval (the time between the onset of disease in the primary infected person and the onset of disease in the secondary infected person). As estimated by Nishiura et al. (2020), He et al. (2020), and Alene et al. (2021), the estimated mean serial interval and the incubation period of COVID-19 are 5.2 and 6.5 days, respectively. Notably, the estimated serial interval is shorter than the estimated incubation period^2^. For seasonal influenza and SARS, the serial interval is longer than the incubation period. This indicates the following important feature of COVID-19—in contrast to seasonal influenza and SARS, a significant number of COVID-19 cases are caused by asymptomatic or pre-symptomatic infection.

Owing to this feature of COVID-19, SARS-CoV-2 is not only transmitted among residents of the same county but also to residents of other counties through cross-county transmission, even before symptom onset. Shang et al. (2021) notes that the epidemiological models based on SIR or SEIR cannot capture this phenomenon^3^. By contrast, the VAR epidemiological model proposed herein does capture this feature by introducing new COVID-19 cases in other counties as a lagged explanatory variable.

Shang et al. (2021) proposed VAR as a promising COVID-19 forecasting model, but the authors did not demonstrate that the predictions made by VAR outperform those of other epidemiological models. The purpose of this study was to show that the county-level prediction of new COVID-19 cases by VAR is superior to that of other epidemiological models.

This paper is organized as follows. Section 2 describes the methodology; Section 2.1 introduces the VAR model, Section 2.2 describes the data, and Section 2.3 describes the estimation. In Section 3, we describe three forecasting scenarios and evaluate other forecasting models. Section 3.1 explains VAR forecasting, Section 3.2 describes the forecast results, and Section 3.3 provides a comparison of our results to those of 11 other forecasting models. We provide a brief conclusion in Section 4.

## 2. METHODOLOGY

In macroeconomics, economic forecasting is important for planning and evaluating government economic policies. In the 1970s, macroeconomists used large-scale models with hundreds of equations to make economic forecasts. However, since Sims (1980) proposed VAR as a new macroeconomic method, no macroeconomist has used such large-scale models. VAR is a multi-equation system in which each variable is a linear function of the past lags of itself and the other variables. The popularity of VAR in economics is owing to its simple forecasting framework ^4^ while outperforming other forecasting frameworks. Here, we show that VAR performs similarly for predicting COVID-19 cases.

### 2.1 Vector autoregression (VAR)

The regular VAR model with *p* lags, denoted by VAR(*p*), can be written as follows:

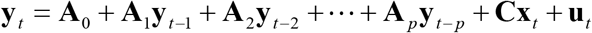

where

***y***_*t*_: *n* × 1 column vector of endogenous variables

***x***_*t*_: *m* × 1 column vector of exogenous variables

***A***_0_: *n* × 1 column vector of constant term

***A***_*i*_: *n* × *n* matrix of lag coefficients to be estimated(*I* = 1,2 …,*p*)

***C***: *n* × *m* matrix of exogenous variable coefficients to be estimated

***u***_*t*_: *n* × 1 column vector of disturbances.

In our model, the column vector is defined as follows:

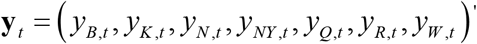

where “ ‘ “ denotes transposition of a vector, *y*_*i*.*t*_ indicates the number of newly confirmed COVID-19 cases in county *i* on day *t*, and B, K, N, NYC, Q, R, and W stand for the New York State counties Bronx, Kings, Nassau, New York City, Queens, Rockland, and Westchester, respectively.

Under the assumption that the time path **y**_*t*_ is stationary^5^, **u**_*t*_ satisfies the following white noise disturbance process:

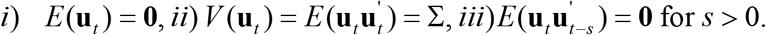

Assumptions *i*) through *iii*) imply that the vector of disturbances is contemporaneously correlated with full rank matrix ∑, but uncorrelated with the leads and lags of the disturbances and uncorrelated with all of the right-hand side variables. Furthermore, each equation is estimated by the ordinary least squares method.

Again, VAR is a multi-equation system in which each variable is a linear function of the past lags of itself and the other variables. Such a framework allows VAR to adequately capture the nature of SARS-CoV-2 transmission at the county level and asymptomatic transmission between counties, which more accurately reflects the cross-county transmission that occurs through the cross-county movement of people.

### 2.2 Data

We analyzed daily new COVID-19 cases in the seven New York State counties assessed by Shang et al. (2021) (Bronx, Kings, Nassau, New York City, Queens, Rockland, and Westchester). Bronx, Kings, and Queens are regarded as regions adjacent to New York City, and this area is classified as a “large central metro” by the Centers for Disease Control and Prevention (CDC). By contrast, Nassau, Westchester, and Rockland have fewer direct connections to New York City; these counties are classified as a “large fringe metro” by the CDC. The data used in this study were downloaded from the following website: US COVID-19 cases and deaths by state | USAFacts^6^. The number of COVID-19 cases reflects the daily cumulative values for each county from March 1, 2020, through August 8, 2021. Based on the accumulated daily counts by county, the daily number of newly infected individuals was calculated by taking the difference. We used newly infected individuals from the county-level daily data for our estimations. There were some days when the number of new cases was recorded as zero, such as February 6 and 26, and March 12, 2021. The reason why the number of new cases was marked as zero is probably due to a delay in recording. It is assumed that the actual number of new cases on these days was added into the new cases of the next day. Therefore, the number of new cases on days with zero new cases was assumed to be half of the number of new cases on the following day. For all days with zero new cases, we took half of the next day’s value as the number of new cases.

To investigate the stationarity of the data, the augmented Dickey–Fuller unit root test^7^ was employed to the new case data of each county. We found that all of the level series had a unit root and were integrated of order one, denoted by I(1)^8^. Consequently, the time path{**y**_*t*_} was concluded to be nonstationary.

### 2.3 Estimation

We used the popular econometric package EViews 12^9^ from IHS Markit. First, we determined the lag order of the VAR model based on the VAR system information criteria, which were the Akaike information criterion (AIC), the Schwarz information criterion (SC), and the Hanna–Quinn information criterion (HQ). The formulae for calculating the AIC, SC, and HQ are defined as (1) through (3) below:

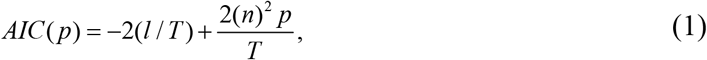

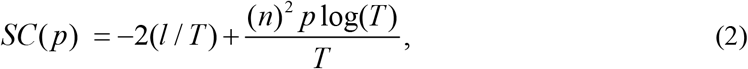

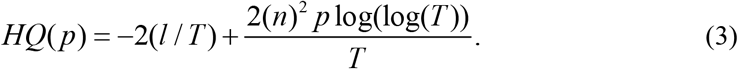

where *n* is the number of explanatory variables, *p* is the lag length, *T* is the sample size, and *l* is the value of the log of the system likelihood function with (*n*)^2^ *p* parameters estimated using *T* observations. The information criteria were calculated with a maximum lag length of 14. AIC is the most commonly used criterion. However, because the sample size (*T*) was large (greater than 500), the AIC defined by (1) did not properly select the lag order. Thus, we applied the SC or the HQ. The SC recommended a lag length of 3, while HQ recommended a lag length of 8. The test results are reported in Appendix Table 1. According to Alene et al. (2021), the estimated average serial interval is 5.2 days (95% CI: 4.9–5.5), which was estimated based on the data of individual infector–infectee pairs. However, the number of new COVID-19 cases was aggregated at the county level, and specific infector–infectee pairs were not able to be identified. Because the data were from online reports of confirmed cases, there was a confirmation lag between symptom onset and confirmation of a positive test result. Assuming that this average serial interval held at the county level, and that we could add the average confirmation lag of 3 to 4 days to the 95% CI of the above serial interval, we could thus regard the duration of infection (the infectious period) as 7.9 to 8.5 days. Based on this duration, a lag order of 8 was selected. We established VAR (8) as the benchmark model for forecasting. The number of estimated coefficients was quite large—more than 500 estimated coefficients—which are not reported here.

All of the data had to be stationary for the VAR estimator to work. As we discussed in Section 2.2, the new COVID-19 case data were nonstationary. Therefore, the VAR estimator did not meet consistency and would be biased. The standard way to solve this problem is to take the difference. However, Sims et al. (1990) and Stock (1994) proved the following useful proposition for large samples: regardless of whether the VAR contains an integrated component, the VAR has consistent ordinary least squares estimators in large samples. Because our sample size was large (greater than 500), the above proposition held for our estimation. In other words, the standard VAR model could be directly applied to estimate the number of new COVID-19 cases by county. Therefore, there was no need to transform the model to a stationary form by differencing.

## 3. FORECASTING

### 3.1 VAR forecasting

As described in Section 2.3, the VAR estimators were consistent in the large sample. Therefore, we conducted VAR estimation to predict the number of new COVID-19 cases in each county. To make comparisons with the other forecasting models, we performed 4-week-ahead forecasting for three scenarios. The VAR (8) model was estimated based on a daily sample, and dynamic forecasting was performed for an out-of-sample period starting on the first forecast day.

1. **6_28 forecast:** Estimate VAR (8) from March 1, 2020, through June 27, 2021, then conduct the 4-week-ahead forecast for June 28, 2021, through July 24, 2021.
2. **7_05 forecast:** Estimate VAR (8) from March 1, 2020, through July 4, 2021, then conduct the 4-week-ahead forecast for July 5 through July 31.
3. **7_12 forecast:** Estimate VAR (8) from March 1, 2020, through July 11, 2021, then conduct the 4-week-ahead forecast for July 12 through August 8.

### 3.2 Results

The root mean square error (RMSE) and the mean absolute percentage error (MAPE) for each of the above three scenarios are reported in Panel (a) and Panel (b) of Appendix Table 2. The RMSE and the MAPE are defined as (4) and (5) below:

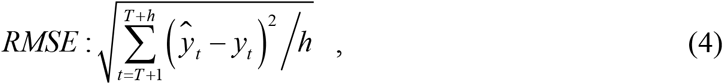

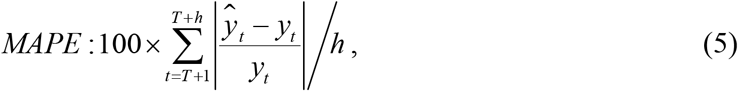

where ŷ_*t*_ is a predicted value and *y*_*t*_ is the real value at time *t*.

Notably, the MAPE values for Rockland and Westchester were larger than the MAPE values for the other counties (Bronx, Kings, Nassau, NYC, and Queens) in all of the scenarios. The latter counties are classified as large central metro communities in the National Center for Health Statistics urban/rural CDC classification, while Rockland and Westchester are classified as large fringe metro communities. The number of new infections was lower in the fringe metro counties of Rockland and Westchester than in the central metro counties. Thus, a shock in the number of new infections is amplified in the fringe metro counties; because the VAR model is linear, it failed to capture such nonlinear shocks. In fact, the regression of the VAR using log-transformed data gave better predictions, even for Rockland and Westchester. However, it performed poorly for the central metro counties. Therefore, the regression of the VAR was done as a level series.

### 3.3 Comparisons

As an example, for the Scenario A) 6_28 forecast, the point forecasts at four specific days, July 3, July 10, July 17, and July 24, were compared with those of 11 other recently proposed forecast models (listed in Appendix Table 3). The point predictions of Scenario B) and C) for these four days were also compared with the same four-point predictions of the other models. We used these four reported point estimates to make comparisons between models. The forecast for July 3 represents a 1-week-ahead forecast based on the data obtained up to June 28. Similarly, the forecast for July 10 represents a 2-week-ahead forecast based on data obtained up to June 28. The same interpretation applies to July 17 (a 3-week-ahead forecast) and July 24 (a 4-week-ahead forecast). The county-level forecasts for the 11 models were extracted from the following CDC files: 2021-06-28-all-forecasted-cases-model-data.cvs, 2021-07-05-all-forecasted-cases-model-data.cvs, and 2021-07-12-all-forecasted-cases-model-data.cvs ^10^. To compare the forecast accuracy between models, the RMSEs of the four-point forecasts are reported in Panel (a) through Panel (g) in Appendix Figure 1.

The results indicated that, compared with the other models, the VAR (8) model exhibited a much better forecasting performance for the 6_28, 7_05, and 7_12 forecasts for Bronx, Kings, Nassau, New York City, and Queens, but not for Rockland and Westchester. For the latter two counties, the forecasting results were comparable with those of the other models. Although not reported here, the mean absolute error and the mean absolute percentage error, which are other forecast error measures, also indicated similar results.

## 4. CONCLUSION

As shown in Section 3, the VAR prediction outperformed the predictions of other state-of-the-art models. The reason for this is that the VAR prediction adequately captures the pre-symptomatic and asymptomatic transmissibility of COVID-19 by introducing data from other counties as lagged explanatory variables. Thus, we strongly recommend the simple VAR model as a framework to accurately predict the regional transmission of COVID-19.

## Data Availability

All data produced in the present study are available upon reasonable request to the authors.

https://usafacts.org/visualizations/coronavirus-covid-19-spread-map

## APPENDIX

**TABLE 1:** LAG-ORDER TEST.

| Lag | AIC | SC | HQ |
| --- | --- | --- | --- |
| 0 | 86.34921 | 86.40786 | 86.37221 |
| 1 | 81.48394 | 81.95311 | 81.66798 |
| 2 | 80.94121 | 81.82091 | 81.28628 |
| 3 | 80.39166 | 81.68189* | 80.89777 |
| 4 | 80.05987 | 81.76063 | 80.72701 |
| 5 | 79.80185 | 81.91314 | 80.63004 |
| 6 | 79.52028 | 82.0421 | 80.5095 |
| 7 | 79.16858 | 82.10092 | 80.31883 |
| 8 | 78.96539 | 82.30827 | 80.27668* |
| 9 | 78.84535 | 82.59875 | 80.31768 |
| 10 | 78.68297 | 82.8469 | 80.31634 |
| 11 | 78.54261 | 83.11707 | 80.33701 |
| 12 | 78.37722 | 83.3622 | 80.33265 |
| 13 | 78.23603 | 83.63154 | 80.3525 |
| 14 | 78.13154* | 83.93758 | 80.40905 |

\* indicates lag order selected by the criterion

**TABLE 2:**
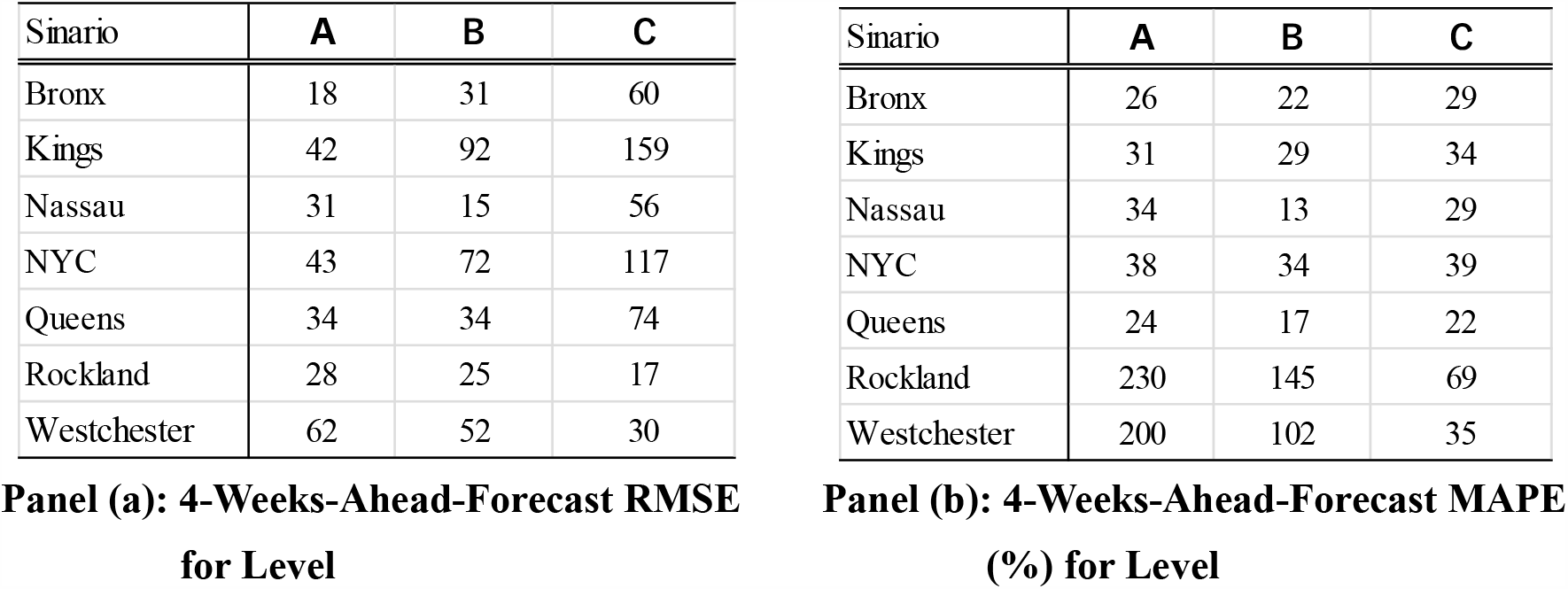
4-WEEK-AHEAD-FORECAST ERROR.

**TABLE 3:** MODEL DESCRIPTIONS.

| Model Name |  | Methods | Classification |
| --- | --- | --- | --- |
| **Var_Lag8 Model** | Kitaoka &Takahashi | Vector Autoregression with 8 lags | Time_series |
| **CMU** | Carnegie Mellon U. | Autoregressive time series model | Time_series |
| **Columbia** | Columbia U. | Metapopulation SEIR model | Mechanistic method |
| **Ensemble** | U. of Massachusetts, Amherst | Combination of 4 to 20 models depending on the availability of forecasts for each location. | Mechanistic method |
| **Facebook** | Facebook AI Research | A machine learning model with an auto-regressive model | Deep_learning based |
| **FRBSF-Wilson** | Federal Reserve Bank of San Francisco/ Wilson | A SIR-derived econometric county panel data model with transmission rate assumed to be function of weather and mobility | Mechanistic method |
| **JHU-APL** | Johns Hopkins U., Applied Physics Lab | Metapopulation SEIR model | Mechanistic method |
| **JHU-UNC-Google** | Johns Hopkins U., U. of North Carolina, and Google | An ensemble of two different models: A multiplicative growth model and a curve fitting model. | Deep_learning based |
| **LANL** | Los Alamos National Laboratory | Statistical dynamical growth model accounting for population susceptibility. | Mechanistic method |
| **PandemicCentral** | Pandemic Central | Random forest machine learning model | Deep_learning based |
| **UGA-CEID** | U. of Georgia, Center for the Ecology of Infectious Disease | Statistical random walk model | Time_series |
| **UVA** | U. of Virginia | An ensemble of three different models: An auto-regressive model, a machine learning (long short-memory) model , and a SEIR model. | Deep_learning based |

**FIGURE 1:**
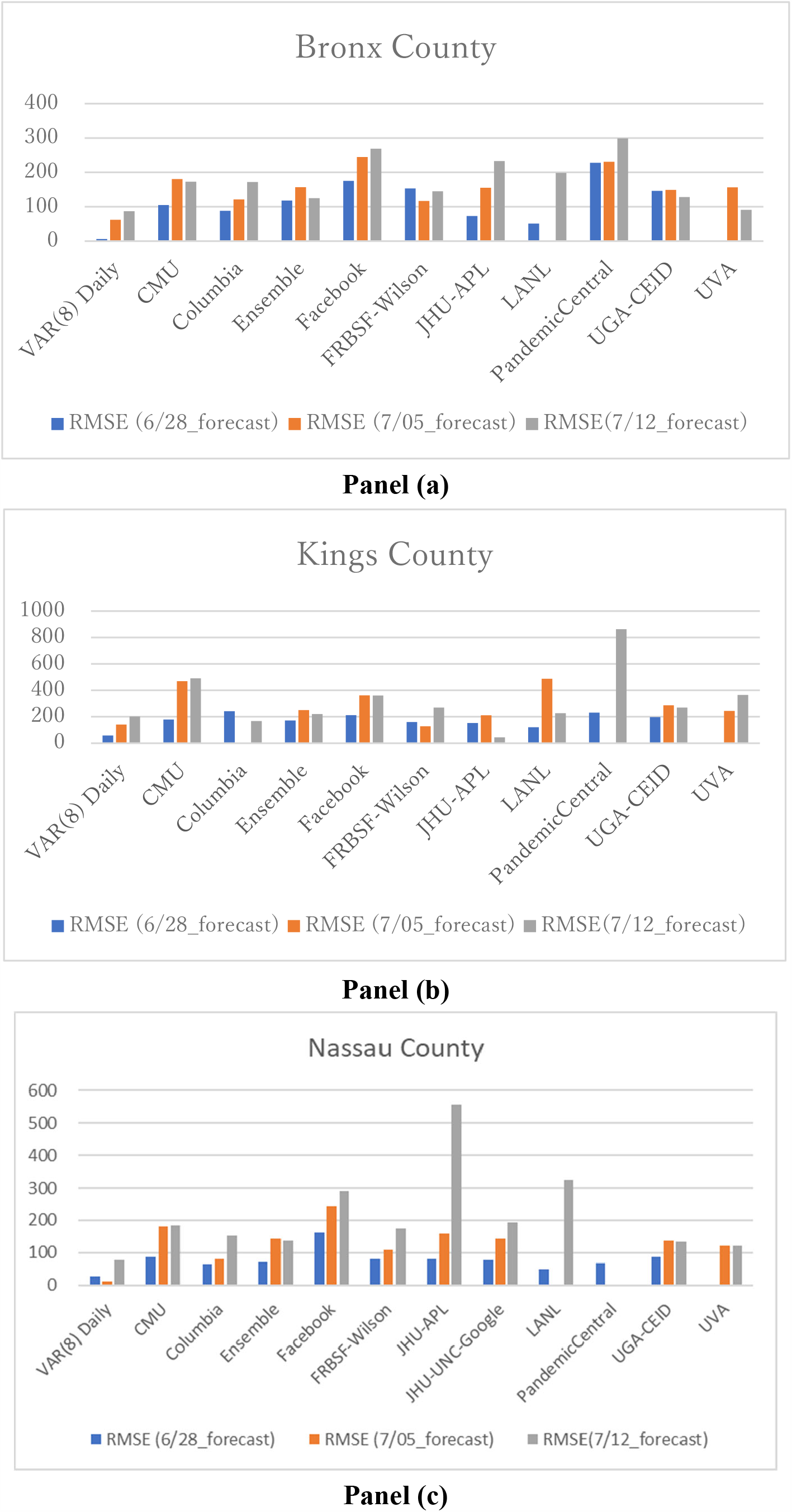
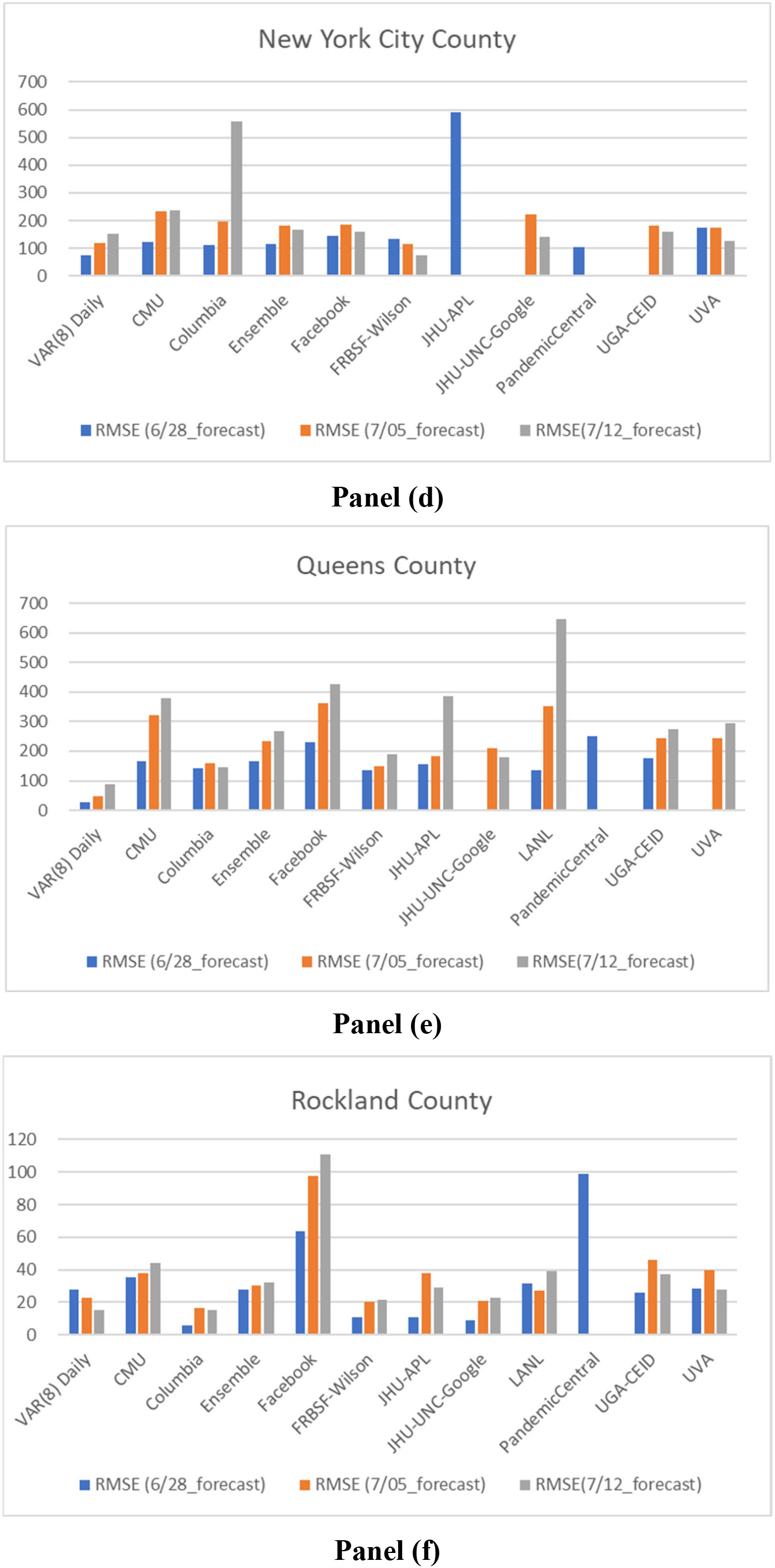
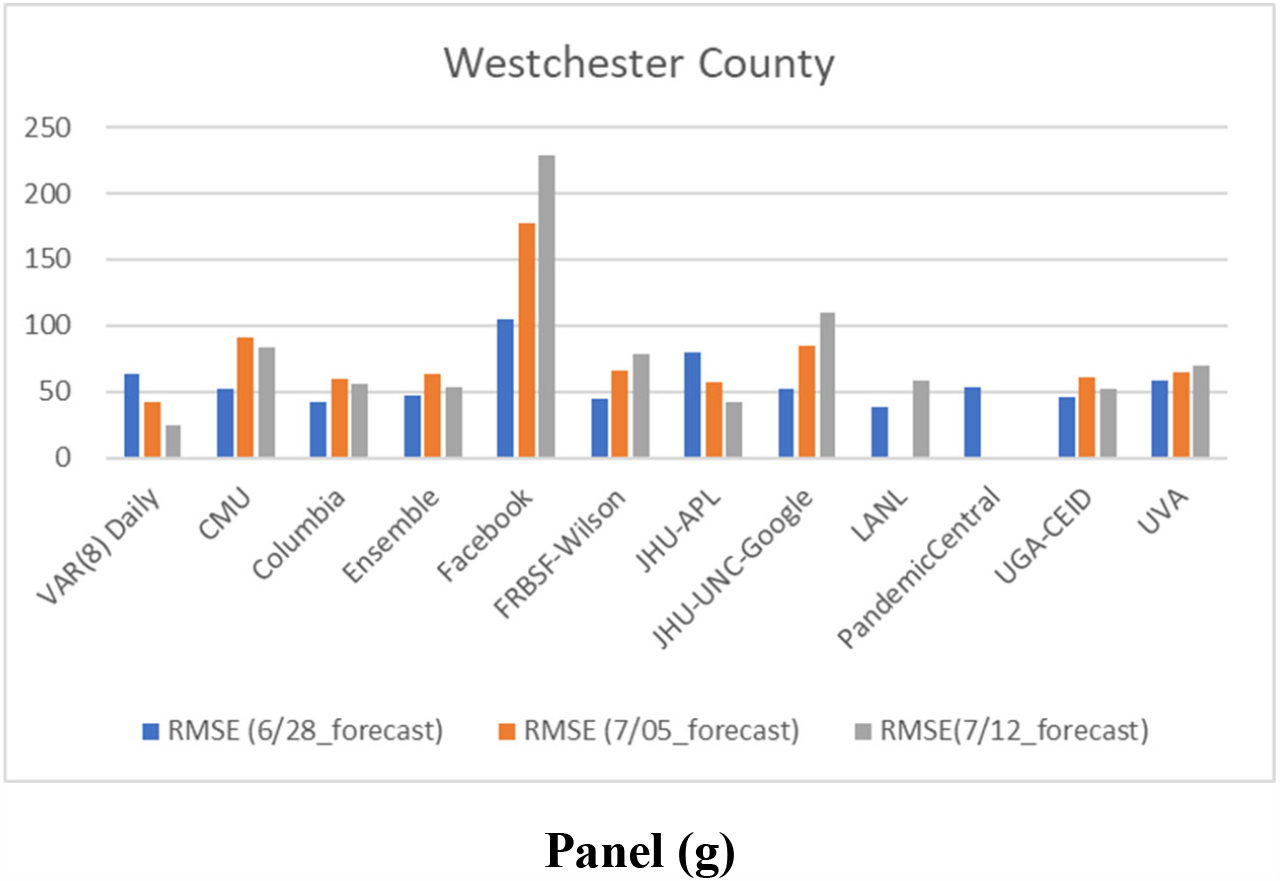
Forecast errors for seven New York counties.

## ACKNOWLEDGMENT

We thank Katherine Thieltges from Edanz (https://jp.edanz.com/ac) for editing a draft of this manuscript.

## DATA AVAILABILITY STATEMENT

The data that support the findings of this research are available in the Excel file in the supplementary material for this article.

## Footnotes

1. Wang et al. (2021) also proposed the VAR model to predict the nationwide daily number of newly COVID-19 cases in the United States. Contrastingly, they used variables potentially correlated to the number of COVID-19 positive cases such as average temperature, precipitation, wind speed, humidity, population density and so on.

2. Note that the serial interval can be negative if a person becomes infected before symptoms appear in the individual who infected them, that is, if the infected person develops symptoms before the person that infected them does.

3. Some forecasting models attempt to incorporate the mobility behavior of individuals into the SRI-based model using a deep learning-based approach.

4. For a comprehensive introduction to VAR estimation, Stock and Watson (2007) is recommended.

5. See, in detail, Key Concept 14.5 in Stock and Watson (2007).

6. County-level data was confirmed by referencing state and local agencies.

7. See, in detail, Key Concept 14.8 in Stock and Watson (2007).

8. If *y*_*it*_ is nonstationary and the first difference of *y*_*it*_, Δ*y*_*it*_, is stationary, then *y*_*it*_ is the integrated one process, denoted by I(1).

9. In addition to EViews, Estima’s RATS is a well-known econometric package specialized for time series analysis.

10. Downloadable from: <u>Previous COVID-19 Forecasts: Cases | CDC</u>.

